## Supplementary material for "Transfer learning for non-image data in clinical research: a scoping review": Data Analysis Code

*Disclaimer: the calculations and numbers are in the focus of this document, and therefore, the text has not been edited at the same level of detail as the corresponding manuscript.*

```
sessionInfo()
```

```
## R version 4.1.0 (2021-05-18)
## Platform: x86_64-apple-darwin17.0 (64-bit)
## Running under: macOS Big Sur 10.16
##
## Matrix products: default
## BLAS:   /Library/Frameworks/R.framework/Versions/4.1/Resources/lib/libRblas.dylib
## LAPACK: /Library/Frameworks/R.framework/Versions/4.1/Resources/lib/libRlapack.dylib
##
## locale:
## [1] en_US.UTF-8/en_US.UTF-8/en_US.UTF-8/C/en_US.UTF-8/en_US.UTF-8
##
## attached base packages:
## [1] stats     graphics  grDevices utils     datasets  methods   base
##
## other attached packages:
## [1] riverplot_0.10 zoo_1.8-9      purrr_0.3.4    stringr_1.4.0  readxl_1.3.1
##
## loaded via a namespace (and not attached):
##  [1] Rcpp_1.0.7         knitr_1.33         magrittr_2.0.1     lattice_0.20-44
##  [5] rlang_0.4.11       fansi_0.5.0        tools_4.1.0        grid_4.1.0
##  [9] xfun_0.24          utf8_1.2.2         htmltools_0.5.1.1  ellipsis_0.3.2
## [13] yaml_2.2.1         digest_0.6.27      tibble_3.1.3       lifecycle_1.0.0
## [17] crayon_1.4.1       RColorBrewer_1.1-2 vctrs_0.3.8        evaluate_0.14
## [21] rmarkdown_2.9      stringi_1.7.3      compiler_4.1.0     pillar_1.6.2
## [25] cellranger_1.1.0   pkgconfig_2.0.3
```

*The document was compiled at 2021-09-30 11:32:19.*

#### Research questions defined in the scoping review protocol

The results are discussed in a different order in the manuscript.

#### To what extent is transfer learning (TL) used in clinical research?

```
n_articles <- nrow(data)
n_articles
```

```
## [1] 83
```

We have identified 83 articles.

#### What is transfer learning used for? (e.g. improve predictions or make them feasible)

The studies' aims have been extracted from a clinical point of view. The frequencies of some common terms are shown below. Note that the aims have been written by the review team and therefore might reflect their word use preferences, although we aimed to be consistent with the specific terms used in the given articles.

```
aim_list <- c('prediction','detection','classification','identification',
              'recognition','diagnosis')

aim_n <- aim_list %>%
  map_int(~ table(str_detect(data$study_purpose, regex(.x, ignore_case = TRUE)))[2])

df <- data.frame(word=aim_list, n=aim_n)

print(df)
```

```
##             word  n
## 1     prediction 23
## 2      detection 20
## 3 classification 17
## 4 identification  2
## 5    recognition  2
## 6      diagnosis  2
```

```
# Sleep staging
sleep_n <- sum(str_detect(data$study_purpose, regex('Sleep staging')))
sleep_n
```

```
## [1] 9
```

The most common specific aim was 'sleep staging' with 9 articles.

The complete list of study aims are shown below.

```
# individual study aims
data$study_purpose
```

```
##  [1] "Classification of resting needle electromyography discharges"
##  [2] "Classification of lung diseases"
##  [3] "Prediction of Alzheimer's dementia"
```

```
##  [4] "Classification of heart sound"
##  [5] "Detection of obstructive sleep apnea"
##  [6] "Diagnosis of COVID-19"
##  [7] "Classification of breathing sound"
##  [8] "Diagnosis of Alzheimer's disease"
##  [9] "Classification of phonocardiogram"
## [10] "Prediction of Alzheimer's Disease"
## [11] "Prediction of surgical complications"
## [12] "Classification of cancers"
## [13] "Prediction of cancer survival"
## [14] "Prediction of survival"
## [15] "Prediction of neurodevelopmental outcomes"
## [16] "Prediction of cancer survival"
## [17] "Classification of mass spectrometry data"
## [18] "Prediction of various clinical outcomes (methodological study)"
## [19] "Prediction of mortality and readmissions"
## [20] "Recognition of breast cancer tissue"
## [21] "Prediction of septic shock"
## [22] "Prediction of peak expiratory flow rate"
## [23] "Classification of drug resistance"
## [24] "Prediction of cancer type"
## [25] "Prediction of patient health at the ICU"
## [26] "Identification of psychiatric stressors"
## [27] "Prediction of diseases"
## [28] "Risk assessment of suicide and self-harm"
## [29] "Prediction of mortality and morbidity"
## [30] "Screening for psychiatric diseases"
## [31] "Prediction of radiation oncology incidents"
## [32] "Detecting self-reports of prescription medication abuse"
## [33] "Arrhythmia detection"
## [34] "Sleep staging"
## [35] "Recognition of stereotypical motor movement"
## [36] "Sleep staging"
## [37] "Detection of diabetes"
## [38] "Detection of epileptic seizure"
## [39] "Identification of atrial fibrillation"
## [40] "Estimation of heart rate"
## [41] "Classification of CVD"
## [42] "Detection of atrial fibrillation"
## [43] "Classification of epileptic seizure"
## [44] "ECG monitoring"
## [45] "Prediction of epileptic seizure"
## [46] "Classification of alcoholism pre-disposition"
## [47] "Detection of neurodevelopmental disorders"
## [48] "Classification of epileptic states"
## [49] "Detection of sleep-disordered breathing"
## [50] "Sleep staging"
## [51] "Detection of seizure"
## [52] "Detection of atrial fibrillation"
## [53] "Detection of atrial fibrillation"
## [54] "Detection of seizure"
## [55] "Sleep staging"
## [56] "Detection of cardiac rhythm"
## [57] "Prediction of dengue fever outbreaks"
```

```
## [58] "Sleep staging"
## [59] "Sleep staging"
## [60] "Detection of spatial gastric slow wave abnormalities"
## [61] "Detection of epileptic seizure"
## [62] "Prediction of glucose levels"
## [63] "Classification of seizure type"
## [64] "Classification of falls risk"
## [65] "Classification of aortic stenosis"
## [66] "Detection of schizophrenia"
## [67] "Triage patients with infectious diseases"
## [68] "Sleep staging"
## [69] "Prediction of gastric cancer morbidity"
## [70] "Sleep staging"
## [71] "Analysis of ECG"
## [72] "Prediction of COVID-19 pandemic"
## [73] "Prediction of glucose levels"
## [74] "Detection of mild cognitive impairment"
## [75] "Detection of seizure"
## [76] "Detection of epileptic seizures"
## [77] "Prediction of various outcomes based on ECG"
## [78] "Classification of atrial fibrillation"
## [79] "Sleep staging"
## [80] "Detection of rare genetic heart disease"
## [81] "Detection of peripheral arterial disease"
## [82] "Estimation of Parkinson's disease severity"
## [83] "Classification of arrhythmia"
```

It is more difficult to summarize (at least quantitatively) the specific reason of using TL. As 4 out of 5 studies compared the performance of the TL model to a non-TL solution, performance improvement seems to be the #1 reason. Also, we observed many examples where the target dataset were very small (sometimes subsets of the source data), limiting the utility of ML methods without the use of transfer learning. In the most extreme cases, the target dataset included data from a specific individual, aiming to develop individualized models, but still utilising data from the whole study population. Similarly, instead of zooming into individuals, other studies utilised data from a disease group (e.g. cancer) to zoom into a specific one in that group (e.g. lung cancer).

#### In which areas of clinical research is transfer learning used?

Medical field was assigned by the reviewers during data extraction. Often more fields were noted in the first place and then the best matching one was chosen in consensus by the reviewers.

```
sort(table(data$study_field), decreasing = T)
```

```
##
##          Neurology          Cardiology            Genetics Infectious diseases
##                 26                  18                   5                   5
##         Psychiatry       Endocrinology        Epidemiology           Pathology
##                  5                   3                   3                   3
##        Pulmonology      Intensive care          Neonatology Otorhinolaryngology
##                  3                   2                   2                   2
##       Pharmacology    Gastroenterology          Geriatrics            Oncology
##                  2                   1                   1                   1
```

```
##             Surgery
##                   1
```

```r
#EEG
eeg_n <- sum(data$extra_eeg=='Yes')
eeg_n
```

```
## [1] 20
```

```r
#ECG
ecg_n <- sum(data$extra_ecg=='Yes')
ecg_n
```

```
## [1] 19
```

Two types of studies stood out with very high frequencies: analysis of EEG (n=20) and ECG (n=19) data.

#### Who uses transfer learning? (computer scientists/clinicians/together)

The authors affiliations were considered in this analysis as a proxy for their professional background, as this information is available in all articles. Technical affiliations included institutions in computer science, engineering, mathematics or similar, while clinical affiliations were defined by ones from medicine, health or similar. Frequencies are shown below.

```r
author_freq <- as.matrix(table(data$authors_clin, data$authors_tech))
names(dimnames(author_freq)) <- c("clinical", "technical")
author_freq
```

```
##         technical
## clinical No Yes
##      No   0  29
##      Yes  4  50
```

```r
# in percentages
author_perc <- round(100*prop.table(table(data$authors_clin, data$authors_tech)))
author_perc
```

```
##
##       No Yes
##   No   0  35
##   Yes  5  60
```

Three out of five studies (60%) were conducted in collaboration between authors from both clinical and technical backgrounds. Studies conducted by authors only from a technical background (35%) were more common than those from only a clinical background (5%).

#### Where are the results published? (clinical/interdisciplinary journals)

```r
sort(table(data$pub_journal), decreasing = T)
```

```
##
##                                 Annu Int Conf IEEE Eng Med Biol Soc
##                                                                   9
##                                        IEEE J Biomed Health Inform
##                                                                   4
##                                                       Physiol Meas
##                                                                   4
##                                                            Sci Rep
##                                                                   4
##                                          BMC Med Inform Decis Mak
##                                                                   3
##                                                        Nat Commun
##                                                                   3
##                                                   Comput Biol Med
##                                                                   2
##                                   Comput Methods Programs Biomed
##                                                                   2
##                                                         PLoS One
##                                                                   2
##                                                             Sleep
##                                                                   2
##                                   AMIA Jt Summits Transl Sci Proc
##                                                                   1
##                                                     Ann Emerg Med
##                                                                   1
##                          Biocybernetics and Biomedical Engineering
##                                                                   1
##                                                    Bioinformatics
##                                                                   1
##                                                 Biomed Eng Online
##                                                                   1
##                          Biomedical Physics and Engineering Express
##                                                                   1
##                            Biomedical Signal Processing and Control
##                                                                   1
##                                                Biosensors (Basel)
##                                                                   1
##                                                BMC Bioinformatics
##                                                                   1
##                                                 Clin Neurophysiol
##                                                                   1
##                                              Comput Intell Neurosci
##                                                                   1
##                                            Comput Math Methods Med
##                                                                   1
##                                              Diabetes Technol Ther
##                                                                   1
##                                                        Eur Neurol
##                                                                   1
##                                              Front Aging Neurosci
##                                                                   1
```

```
##                                                        Front Digit Health
##                                                                         1
##                                                        Front Hum Neurosci
##                                                                         1
##                                                              Front Neurol
##                                                                         1
##                                                          Front Psychiatry
##                                                                         1
##                                                             Front Psychol
##                                                                         1
##                                                      Health Inf Sci Syst
##                                                                         1
##                                                       Healthc Inform Res
##                                                                         1
##                                                       Healthcare (Basel)
##                                                                         1
##                                           IEEE Trans Biomed Circuits Syst
##                                                                         1
##                                                     IEEE Trans Biomed Eng
##                                                                         1
##                                         IEEE Trans Neural Netw Learn Syst
##                                                                         1
## IEEE Transactions on Neural Systems and Rehabilitation Engineering
##                                                                         1
##                                                      Inform Med Unlocked
##                                                                         1
##                                          Informatics in Medicine Unlocked
##                                                                         1
##                                           Int J Comput Assist Radiol Surg
##                                                                         1
##                                           Int J Environ Res Public Health
##                                                                         1
##                                                       Int J Neural Syst
##                                                                         1
##                                                                      IRBM
##                                                                         1
##                                                         J Alzheimers Dis
##                                                                         1
##                                                          J Biomed Inform
##                                                                         1
##                                                             J Biomed Res
##                                                                         1
##                                                      J Healthc Inform Res
##                                                                         1
##                                                       J Med Internet Res
##                                                                         1
##                                                             J Neural Eng
##                                                                         1
##                                                       J Neurosci Methods
##                                                                         1
##                        Journal of Medical and Biological Engineering
##                                                                         1
##                    Journal of Medical Imaging and Health Informatics
##                                                                         1
```

```
##                                                  Neural Netw
##                                                            1
##                                               NPJ Digit Med
##                                                            1
##                                            PeerJ Comput Sci
##                                                            1
##                                            Phys Eng Sci Med
##                                                            1
##                                    Proc Natl Acad Sci U S A
##                                                            1
##                                             Sensors (Basel)
##                                                            1
```

```
# IEEE journals
is_IEEE_n <- sum(str_detect(data$pub_journal, regex("IEEE", ignore_case = TRUE)))
is_IEEE_n
```

```
## [1] 17
```

The overall picture is that the majority of articles were published in interdisciplinary journals with a significant technical focus, despite that we considered only medical databases (PubMed, EMBASE, CINAHL). One in five articles (n=17) were published in IEEE-related journals (or proceedings). There are almost no clinical mainstream journals in the list, but a few general scientific journals (Sci Rep, Nat Commun, Plos One) with a broader audience.

##### What type of non-image data is transfer learning applied for? (e.g. tabular, time series, text, voice)

```
type_freq <- table(data$data_type_target_domain)
type_freq
```

```
##
##       Audio     Tabular        Text Time series
##          10          15           7          51
```

```
round(100*prop.table(type_freq))
```

```
##
##       Audio     Tabular        Text Time series
##          12          18           8          61
```

```
# different source and target types
type_diff_n <- sum(data$data_type_source_diff=='Yes')
type_diff_n
```

```
## [1] 36
```

```r
# difference in data type by target domain
table(data$data_type_target_domain, data$data_type_source_diff)
```

```
##
##              No Yes
##   Audio        1   9
##   Tabular     13   2
##   Text         7   0
##   Time series 26  25
```

```r
# use of image models
table(data$data_type_source_image)
```

```
##
##  0  1
## 50 33
```

The most common target data type was time series (n=51), then tabular data (n=15), audio (n=10) and text (n=7).

```r
# cross-tab source and target
cross_source_target <- table(data$data_type_source_combined, data$data_type_target_domain)
cross_source_target
```

```
##
##                     Audio Tabular Text Time series
##   Audio                 1       0    0           0
##   Audio, image          1       0    0           0
##   Image                 5       2    0          23
##   Tabular               0      13    0           0
##   Text                  2       0    7           0
##   Time series           1       0    0          26
##   Time series, image    0       0    0           2
```

```r
# if a study used different two different types of models, then these contribute
# to both types
# make order: text, audio, time series, tabular, image
perm_col <- c(3, 1, 4, 2)
cross_source_target_simple0 <-
  rbind(cross_source_target[5,perm_col],
        cross_source_target[1,perm_col]+cross_source_target[2,perm_col],
        cross_source_target[6,perm_col]+cross_source_target[7,perm_col],
        cross_source_target[4,perm_col],
        cross_source_target[2,perm_col] +
          cross_source_target[3,perm_col] +
          cross_source_target[7,perm_col])

# add a column of zeros to represent image target types
cross_source_target_simple <- cbind(cross_source_target_simple0, rep(0,5))
```

We were surprised seeing many studies (n=36) reusing models from a different source data type than the target itself. The frequencies of different combinations are shown in the last table above, where the rows represent the source domain and the columns represent the target domain. Three studies included and compared source models of different types, that's why multiple types appear in some rows. These studies contributed with two records to the Sankey diagram.

```
cols5 <- c(rgb(0, 158, 115, 255, maxColorValue = 255), # bluish green
           rgb(86,180,233,maxColorValue = 255), # sky blue,
           rgb(0, 0, 128, 255, maxColorValue = 255), # navy
           rgb(255, 140, 0, 255, maxColorValue = 255), # dark orange
           rgb(178, 34, 34, 255, maxColorValue = 255) # firebrick
)



nodesX <- data.frame( ID= c(paste0("B",1:5), paste0("F",1:5)),
                     x= c(rep(2,5),rep(1,5)),
                     col=c(cols5,cols5),
                     labels= c('Text','Audio','Time series','Tabular','Image',
                               'Text','Audio','Time series','Tabular',''),
                     stringsAsFactors= FALSE )

edgesX <- data.frame(
    N1 = paste0("B",rep(1:5,each=5)),
    N2 = paste0("F",rep(1:5,5)),
    Value = as.vector(t(cross_source_target_simple)),
    col = rep(cols5,each=5)
)

rX <- makeRiver(nodesX, edgesX)

d <- list(srt=0, textcex=0.8, textcol = 'white')
riverplot(rX, plot_area=c(1,1), nodewidth=7, default_style=d,node_margin=0.025,
            edgestyle='sin', nsteps = 1000)
```

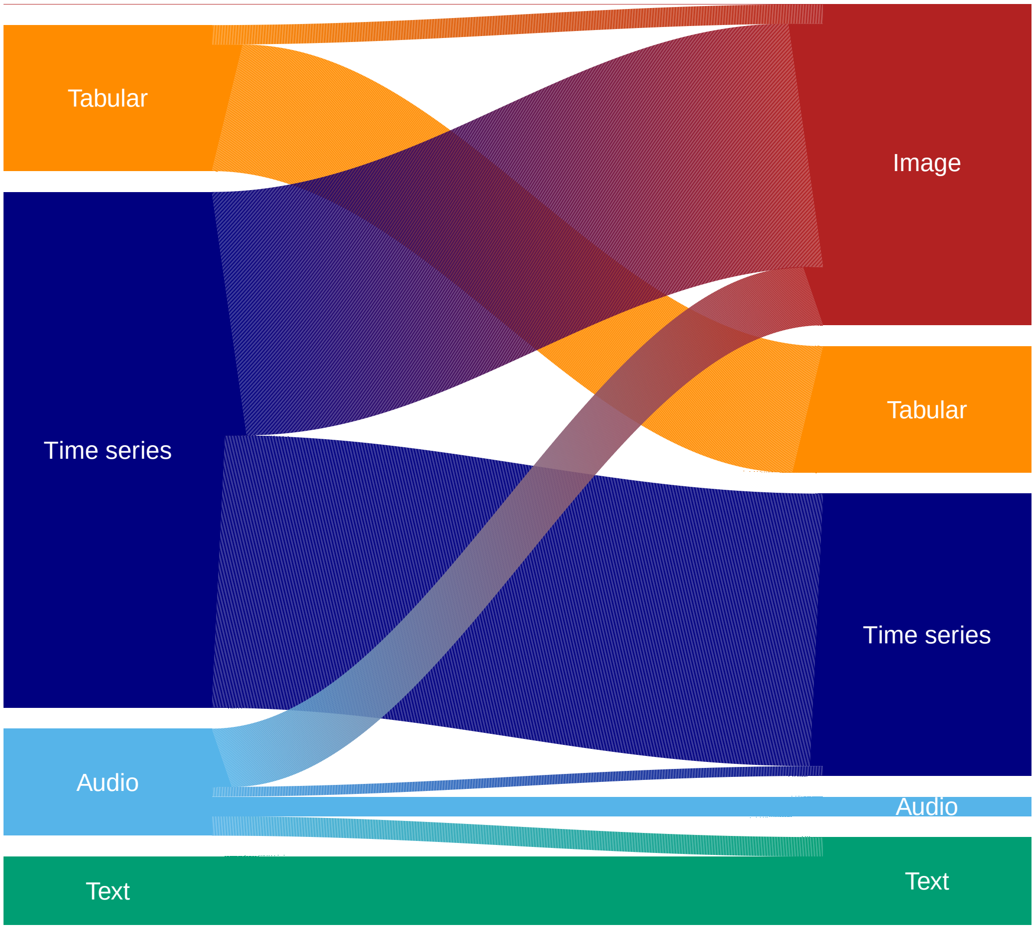


The riverplot package has a well-documented issue (see p. 13 in the package documentation) i.e. displaying vertical white lines, that we tried to overcome by increasing the number of interpolating segments. This led to unwated triangles both on the left and the right sides, which are covered by rectangles of a matching color (done outside of R) in the version used in the manuscript.

#### What are the sizes of the datasets used in transfer learning (both in the source and target domains)?

Differences in units made it unfeasible to examine this question. One of the main contributing factors to this was that source and target data types were different more often than we expected (n=36). E.g. audio recordings in minutes, number of pictures, tweets.

```r
# difference
table(data$data_type_source_diff)
```

```
##
##  No Yes
##  47  36
```

##### What kind of models are reused with transfer learning? (e.g. CNN, LSTM)

As expected, neural network-based methods dominated the type of reused models, with a very few exceptions (n=3) reusing traditional statistical models or SVMs. The frequencies are shown below (the numbers don't necessarily add up to the number of articles, because some studies used multiple methods and/or their combinatation).

```r
# CNN
table(data$TLModel_cnn)
```

```
##
##  0  1
## 25 58
```

```r
# RNN
table(data$TLModel_rnn)
```

```
##
##  0  1
## 65 18
```

```r
# Vanilla NN
table(data$TLModel_nn)
```

```
##
##  0  1
## 76  7
```

```r
# Transformers
table(data$TLModel_transformers)
```

```
##
##  0  1
## 77  6
```

```r
# Autoencoders
table(data$TLModel_autoencoder)
```

```
##
##  0  1
## 78  5
```

```r
# Traditional statistics
table(data$TLModel_trad_stat)
```

```
##
##  0  1
## 81  2
```

```r
# SVM
table(data$TLModel_svm)
```

```
##
##  0  1
## 82  1
```

```r
TL_type_table <- table(data$type_of_tl)
TL_type_table
```

```
##
##                    Both Feature-representation            Fine-tuning
##                       7                     18                     58
```

Fine-tuning was approx. 3x as common (n=58) as feature representation transfer (n=18). Seven studies used both.

```r
table(data$type_of_feature)
```

```
##
##                                          Boosting, Traditional statistics
##                                                                         1
##                                                                        NN
##                                                                         7
## NN, Classification tree, random forest, SVM, Naive Bayes, K-nearest neighbor
##                                                                         1
##                                     NN, CNN, hierarchical attention network
##                                                                         1
##                                        NN, CNN, RNN, Traditional statistics
##                                                                         1
##                                             NN, SVM, Traditional statistics
##                                                                         1
##                                                                       RNN
##                                                                         2
##                                                                       SVM
##                                                                         8
##                                                                  SVM, kNN
##                                                                         1
##                                                 SVM, Traditional statistics
##                                                                         1
##                                                      Traditional statistics
##                                                                         1
```

```r
# SVM
table(data$FeatureModel_svm)
```

```
##
##  0  1
## 13 12
```

```r
n_svm <- sum(data$FeatureModel_svm,na.rm = T)
# NN
table(data$FeatureModel_nn)
```

```
##
##  0  1
## 14 11
```

```r
n_nn <- sum(data$FeatureModel_nn,na.rm = T)
```

Support vector machine (SVM) was the most common final model (n=12) out of the 25 studies using feature representation transfer, followed closely by 'vanilla' neural networks. Seven studies used and compared multiple models.

##### Are the reused models and implementation code publicly available?

```r
# Model availability and development
n_avail_model <- sum(data$source_model_available=='Yes')
tab_orig_avail <- table(data$origin_of_reused_model,data$source_model_available)
tab_orig_avail
```

```
##
##                    No Yes
##   Both              1   0
##   Current study    38   8
##   External source   3  33
```

```r
# % of current-external within available
round(100*prop.table(tab_orig_avail,2))
```

```
##
##                    No Yes
##   Both              2   0
##   Current study    90  20
##   External source   7  80
```

```r
# Code availability
table(data$code_available)
```

```
##
##  No Yes
##  61  22
```

Half of the studies used an publicly available model (n=41), among which those models dominated that were readily available in the used software packages.

Code availability was low, only one in four studies published their code, most of these were available on GitHub.

##### What software was used to implement transfer learning? (e.g. Python, R, MATLAB)

```
table_software <- table(data$software)
table_software
```

```
##
## MATLAB    N/A Python
##     12     28     43
```

```
n_python <- table_software['Python']
n_matlab <- table_software['MATLAB']
```

Python (n=43) was the dominant software among those who reported this detail, followed by MATLAB (n=12). One in three articles did not report the software they used at all. We did not find a single study using R.

These numbers represent the software used for the analysis and not for data processing. However, if the authors reported software/packages used for data processing, but not for the analysis, then we assumed that they used the same. If packages like PyTorch, tensorflow or keras were mentioned, then we assumed that the authors used Python (their native language), as these packages became available in R only recently.

##### Is the transfer learning solution compared to a (benchmark) solution without transfer learning? If yes, what are the costs or benefits of transfer learning?

```
table(data$tl_compared_to_non_tl)
```

```
##
##  No Yes
##  16  67
```

Four in five studies compared the TL results to a non-TL solution. The majority of these studies included multiple models and performance metrics and therefore it is difficult to quantify the costs/benefits of using TL, however shorter running time/less iteration needed were mentioned several times among the benefits.

##### Trend in using transfer learning? (calendar year)

```
# within 12 months from search (May 18, 2021)
sum(data$pub_epubdate > '2020-05-18')
```

```
## [1] 52
```

```
table(data$pub_year)
```

```
##
## 2012 2018 2019 2020 2021
##    1    5   10   41   26
```

```
data$pub_quarter <- as.yearqtr(data$pub_epubdate, format = "%Y-%m-%d")
freq_Q <- table(data$pub_quarter)
freq_Q
```

```
##
## 2013 Q1 2018 Q1 2018 Q2 2018 Q3 2018 Q4 2019 Q1 2019 Q2 2019 Q3 2019 Q4 2020 Q1
##       1       1       2       2       1       2       1       3       4       9
## 2020 Q2 2020 Q3 2020 Q4 2021 Q1 2021 Q2
##      11       8      14      16       8
```

```
df_quarter <- data.frame(t = seq(2018.125, 2021.125, 0.25),
                         n =  as.numeric(freq_Q[2:14]))

df_quarter_ext <- rbind(df_quarter, c(2021.375, 14.86))
df_quarter_ext$per2 <- df_quarter_ext$t < 2019.5
df_quarter_ext$t2019_6 <- df_quarter_ext$t - 2019.5

model <- lm(n ~ t2019_6 + t2019_6:per2, data = df_quarter_ext)
model_coef <- coef(model)

colRM <- c(rgb(0,171,164,255,maxColorValue = 255), #green
           rgb(250,187,0,255,maxColorValue = 255), #yellow
           rgb(101,90,159,255,maxColorValue = 255), #purple
           rgb(226,0,122,255,maxColorValue = 255)) #magenta
plot(-9,-9,
     xlab = 'Calendar year', ylab = '',
     main = 'Number of articles published',
     xlim = c(2018,2022), ylim = c(0, 20),

     xaxt = 'n', yaxt = 'n',
     bty = 'n')
abline(v = seq(2019,2021,1), col = 'lightgray', lwd = 0.5, lty = 1)
axis(1, at = seq(2018, 2022), pos = 0)
axis(2, at = seq(0,20,5), las = 1, pos = 2018)
x <- seq(2018.125, 2021.375, 0.05)
y <- model_coef[1] + model_coef[2]*(x-2019.5) + model_coef[3]*(x-2019.5)*(x<2019.5)
lines(x, y, lty = 2)
points(df_quarter$t, df_quarter$n, pch = 19,
       col = c(rep(colRM,3), colRM[1]))
points(2021.375, 14.86, pch = 1, col = colRM[2])
legend(2018.1, 20.9, legend=c('Q1', 'Q2', 'Q3', 'Q4'),
       col = colRM, pch = 19,box.lty=0)
```

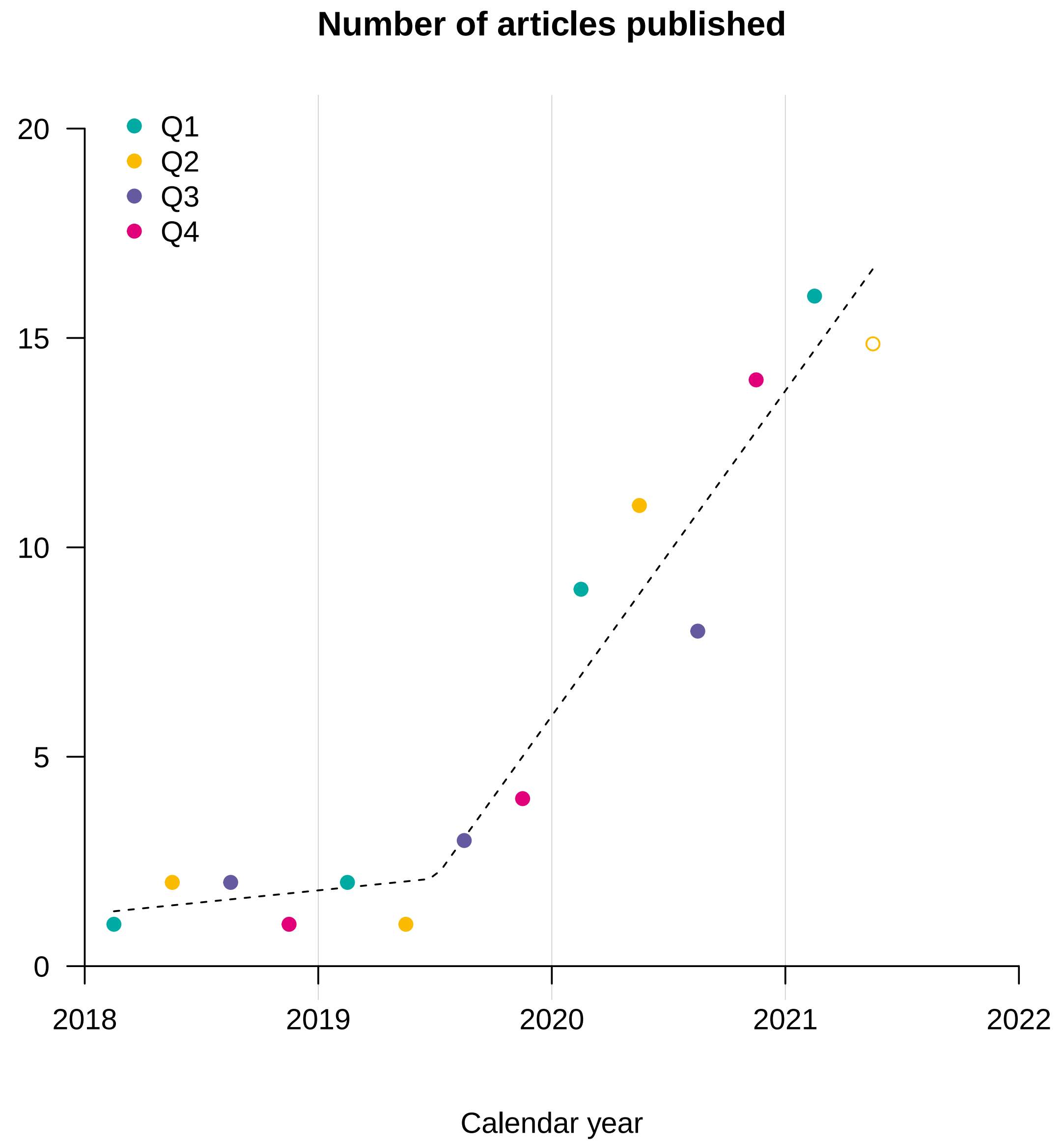


The number of articles seems to increase rapidly from 2019, although the overall number is still quite low

### Questions formulated after publishing the protocol

#### To what extend are open access datasets used?

```
table(data$open_access_dataset)
```

```
##
##  No Yes
```

```
## 28  55
```

```r
# open access - code sharing (reproducability)
table(data$open_access_dataset, data$code_available)
```

```
##
##       No Yes
##   No  22   6
##   Yes 39  16
```

```r
# ImageNet
table(data$extra_imagenet)
```

```
##
##  No Yes
##  56  27
```

```r
# PhysioNet
table(data$extra_physionet)
```

```
##
##  No Yes
##  58  25
```

```r
n_phys <- sum(data$extra_physionet=='Yes')
```

Two out of three articles utilized some open access dataset.

ImageNet-based models seemed to have a large impact on clinical studies even those analyzing non-image data.

The PhysioNet database was a common source for data with 25 studies utilizing data from this repository.

#### Summaries by data types

```r
# transformation
table(data$data_type_target_domain, data$data_type_source_diff)
```

```
##
##               No Yes
##   Audio        1   9
##   Tabular     13   2
##   Text         7   0
##   Time series 26  25
```

```r
# open access
table(data$data_type_target_domain, data$open_access_dataset)
```

```
##
##                No Yes
##   Audio         2   8
##   Tabular       6   9
##   Text          6   1
##   Time series 14  37
```

```r
# ImageNet
table(data$data_type_target_domain, data$extra_imagenet)
```

```
##
##                No Yes
##   Audio         6   4
##   Tabular      14   1
##   Text          7   0
##   Time series 29  22
```

```r
# PhysioNet
table(data$data_type_target_domain,data$extra_physionet)
```

```
##
##                No Yes
##   Audio         8   2
##   Tabular      15   0
##   Text          6   1
##   Time series 29  22
```
